# Wearable Sensors Predict Exertion Without Changes in Pulse Rate for Frail End-Stage Kidney Disease Patients

**DOI:** 10.64898/2026.11.12.26367919

**Authors:** Mikayla E. Deehring, Elizabeth C. Lorenz, H. Philo Katzman, Rebecca Isac, Farzan Sasangohar, Atiya Dhala, Momona Yamagami

## Abstract

End-Stage Kidney Disease (ESKD) patients require careful exertion monitoring during prehabilitation to ensure safety and efficacy. Traditional prehabilitation typically relies on heart rate to estimate physical exertion; however, this is problematic for ESKD patients due to their lower physical capacity and the high prevalence of beta-blocker prescriptions, which prevent typical heart rate responses during exercise. In this study, we investigated whether multi-modal wearable sensors could better predict the Rating of Perceived Exertion (RPE) in (N=13) ESKD patients and (N=13) healthy older adult controls. We collected continuous physiological data—including pulse rate, electrodermal activity, skin temperature, and accelerometer data—using a commercial smartwatch during prehabilitation sessions. Using individualized machine learning pipelines, we compared regression models built solely on pulse rate against comprehensive multi-feature models. We found distinct differences between the groups: baseline pulse-rate models performed substantially worse for the ESKD cohort, whereas they maintained predictive validity for controls. Conversely, multi-feature models significantly improved predictive performance for ESKD participants, achieving a Root Mean Square Error of 1.33. Feature selection confirmed that ESKD models relied heavily on physiological data beyond pulse rate to compensate for the lack of cardiovascular variability. These results demonstrate that heart rate alone is an insufficient biomarker for this population, and that remote exertion monitoring for ESKD patients requires distinct, multi-modal predictive algorithms rather than those designed for healthy adults.

## 1 Introduction

An estimated one in seven adults in the United States live with chronic kidney disease (CKD) [1]. At the endpoint of disease progression, end-stage kidney disease (ESKD), patients require dialysis or a kidney transplant to survive [2]. However, ESKD patients with frailty are less likely to qualify for a kidney transplant or survive the duration of the waitlist [3]. Frailty is generally defined as “… a clinically recognizable state of increased vulnerability, resulting from aging-associated decline in reserve and function across multiple physiologic systems such that the ability to cope with everyday or acute stressors is compromised,” [4]. Beyond acting as a barrier to kidney transplantation, frailty is associated with other severe negative outcomes, including falls, mortality, hospital readmission, and delayed graft function [5]–[7]. This is a critical issue given its high prevalence: 14–72% of hemodialysis patients experience frailty, compared to just 11% of community-dwelling older adults [5], [8].

Targeted physical rehabilitation and increased physical activity have been shown to safely reduce frailty and improve overall physical performance in this population [9]–[11]. Structured nutrition and physical exercise programs prescribed to improve perioperative outcomes of major surgeries can be referred to as prehabilitation [12]. In this work, we consider exercise prehabilitation only. Despite these positive outcomes for CKD patients both before and after kidney transplant, a major barrier to participation is the frequency and length of prehabilitation. Protocols investigated in prior works typically require multiple in-person clinic or intradialytic (i.e., occurring during dialysis) visits each week for several weeks or months [13]– [19]. In addition to these long protocols, patients may need to maintain fitness for years while they wait for a kidney transplant [20], [21]. This frequent and prolonged nature of prehabilitation is burdensome for patients who already attend many appointments, face barriers to accessing care due to disability [22], [23], and experience financial difficulties [22]. Although intradialytic exercise addresses some travel issues, it requires specialized staff who are not available in every clinic. Due to these difficulties, home-based exercise may be a preferable alternative for patients with ESKD.

Although home-based programs address some issues presented by in-clinic programs, they present a major limitation: the difficulty of effectively tracking patient adherence and progress remotely [24], [25]. Tracking improves adherence and is desired by patients [22], [24], [26], but it requires accurate measurement of exercise duration and intensity [27]. Heart rate is frequently used to prescribe and track exercise intensity [28], [29]. However, heart rate alone is often an ineffective metric for frail CKD/ESKD patients; their frailty can cause autonomic dysfunction, making standard age-based estimates of maximum heart rate inaccurate [30], and their frequent use of beta-blockers to control blood pressure prevents typical heart rate increases in response to exertion [31]–[33]. Therefore, self-reported exercise intensity, such as rating of perceived exertion (RPE), may serve as a complementary source for effectively monitoring and prescribing exercise intensity in this population.

RPE is a method in which the person exercising sums up their entire experience of exertion into a single subjective number [34]. Commonly used RPEs include the Borg RPE scale, which ranges from 6 (no exertion) to 20 (maximum exertion) to roughly correlate with heart rate; and the CR10 scale, which ranges from 0 (no exertion) to 10 (maximum exertion) to improve patient interpretability [35], [36]. Despite the benefits and practicality of RPE scales, such ratings are subjective, and they cannot be directly measured by sensors. Consequently, prior works have explored predicting exertion and fatigue using wearable sensors across diverse applications, including athletic performance (e.g., running, cycling, and resistance training) [37]–[45], occupational safety [46], [47], and clinical contexts [48], [49]. However, to our knowledge, no works have focused on predicting exertion in older adults or ESKD patients, and only one work investigates a patient population (multiple sclerosis) [49]. This represents a major gap, as physiological signals in aging and chronic disease populations differ significantly from those of their young, healthy counterparts [50]– [52]. Specifically, in the older patient population, we anticipate that heart rate will be a poor predictor of RPE due to the patients’ low physical capacity and their use of beta-blockers. Furthermore, existing models often rely on sensors ill-suited for the long-term, high-frequency prehabilitation of frail patients, such as cumbersome whole-body inertial measurement unit (IMU) systems [42], [44], [47] or lower-extremity IMUs that fail to capture seated upper-body exercises [39]. There is notably little work using smartwatch data [53], which offers a convenient, accessible alternative that patients may already wear in their daily lives.

In this study, we evaluate the efficacy of multi-modal smartwatch data for predicting RPE in a clinical population compared to a healthy cohort. Using data collected during active prehabilitation sessions from 13 participants with ESKD and 13 controls, all recruited between the ages of 55 and 80, we developed and compared personalized machine learning models using a broad feature set against baseline models that relied solely on pulse rate (PR). Our results demonstrate that PR-only models are insufficient for capturing perceived exertion in ESKD patients, consistently underperforming compared to multi-modal approaches. We also find that pulse rate is less commonly used as a feature, with lower feature importance in models tuned for ESKD patients than in those tuned for controls. These findings underscore the necessity of multimodal sensing in clinical populations where physiological responses to exertion may be atypical, providing a more robust framework for remote exercise monitoring and prescription for ESKD patients with frailty.

## 2 Materials and Methods

### 2.1 Experiments

All experimental procedures and protocols were approved by the Institutional Review Boards (IRB) of both Rice University (IRB-FY2024-244; approved March 6, 2024) and Houston Methodist Hospital (PRO00038084:1 Enabling Access to Prehabilitation for Kidney Transplant Candidates who are Frail; approved August 2, 2024). The portion of the study including ESKD patients is registered as a clinical trial (Prehabilitation for Kidney Transplant Candidates, NCT06886256). Written informed consent was obtained from all participants prior to the start of the study sessions.

#### Sensors and Data Labeling

Each participant was fitted with an Empatica EmbracePlus smartwatch [54] on their dominant wrist. Participants were also fitted with a Polar H10 heart rate monitor [55] unless they reported having an internal pacemaker or defibrillator. At the beginning of study sessions, the Borg CR10 scale [36] was explained to the participant. During the study, participants were asked to report their RPE approximately once per minute. Participants were also instructed to report any changes in RPE, even if they were not asked. Timestamped RPE and exercise type tags were recorded using custom software built using Unity version 2022.3.21f1 [56] on a desktop computer. Exercise types were tagged as aerobic exercise (using a treadmill or a stationary bicycle), resistance exercise (using resistance bands), taking a break between sets, and resting at the beginning of the session, before any exercise occurred.

#### Prehabilitation Protocol

The prehabilitation protocol used in this study was developed by Houston Methodist Hospital. It consists of six resistance exercises using resistance bands and aerobic exercise on a treadmill or stationary bicycle. The six resistance exercises included: chair squats, bent-over rows, chest press, seated leg extensions, hip abductions, and glute bridges. At the completion of the prehabilitation protocol, participants were asked to rest with sensors on for a few minutes for their RPE to return to 1 or below.

#### Control Participants

We recruited 14 participants between the ages of 55–80 without self-reported disabilities or chronic conditions for a single two-hour session at Rice University. One participant was excluded from analysis due to insufficient smartwatch data, and data from the remaining 13 participants were analyzed (mean age 70 ± 6.3 years; 7 women; Table 1).

**Table 1:** Participant Demographics.

|  | **Control**<br>(mean ± std) | **ESKD**<br>(mean ± std) |
|---|---|---|
| FFP Criteria Met | 0 ± 0.6 | 2 ± 0.6 |
| Age (years) | 70 ± 6.3 | 64 ± 5.3 |
| BMI | 24 ± 4.4 | 31 ± 5.5 |
| Height (inches) | 67 ± 4.7 | 64 ± 4.0 |
| Weight (pounds) | 154 ± 32.6 | 180 ± 34.7 |
| Number of Women | 7 | 7 |
| Number Using Beta Blockers | 3<sup>a</sup> | 11 |
| Total Number of Participants | N=13 | N=13 |

**Note:** FFP: Fried Frailty Phenotype; BMI: Body Mass Index.
<sup>a</sup>Two control participants could not be reached to confirm beta-blocker usage.

At the start of the session, participants completed a demographic survey and underwent assessments for the Fried Frailty Phenotype (FFP) [57] (Table 1). The Fried Frailty Phenotype is defined by five components: unintentional weight loss, self-reported exhaustion, weakness (reduced grip strength), slow walking speed, and low physical activity with 1–2 criteria indicating prefrailty and ≥3 criteria indicating frailty [57]. Control participants were then asked to complete 3 sets of 10 repetitions of all 6 resistance exercises with a target RPE of 3–5. After completing these exercises, participants performed an additional 3 sets of 10 chair squats with a heavier resistance band, aiming for an RPE of 7. Participants then walked or jogged on a treadmill for 15 minutes at a target RPE of 3–5, followed by an additional 5 minutes at a target RPE of 7.

#### ESKD Participants

We recruited 15 frail or prefrail (according to FFP [57]) ESKD patients aged 55–80 undergoing evaluation for kidney transplantation to participate in two one-hour sessions at Houston Methodist Hospital. Two participants were excluded from analysis: one due to a data collection error, and one who reported an RPE of 0 for the entire session, which precluded an individualized model. Data from the remaining 13 participants were analyzed (mean age 64 ± 5.3 years; 7 women; Table 1). All participants with ESKD were receiving maintenance dialysis.

During the first session, demographic information and FFP measures were collected (Table 1). During the second session, a licensed physical therapist selected four of the six resistance exercises based on each participant’s capabilities and made any necessary adjustments to accommodate participants’ varying physical capacities for exercise (e.g., seated rows, not using a resistance band, etc.). Participants completed 3 sets of 10 repetitions for each selected exercise with a target RPE of 3–5. After the resistance exercises, participants completed up to 10 minutes of aerobic exercise (recumbent or hand cycling) according to their capabilities. For one participant, a six-minute walk test performed during the second visit replaced cycling due to high exertion levels.

### 2.2 Machine Learning

#### Data Processing

Data were downloaded from the Empatica CareLabs portal [58] and included two categories: digital biomarkers, calculated by Empatica at one-minute intervals, and raw sensor data sampled at higher frequencies. From the digital biomarkers, we used pulse rate and actigraphy counts, while the raw sensor data included triaxial accelerometer data, electrodermal activity (EDA), and skin temperature. To synchronize these data types, the raw sensor data were processed into one-minute windows with a 20-second stride. Pulse rate and actigraphy counts were then interpolated forward to match these windowed timestamps. Feature extraction was performed for each window as follows: accelerometer features were extracted according to Preece et al. [59], and EDA features were extracted using the methods in Taylor et al. [60]. For skin temperature, the mean and standard deviation were calculated for each window. SciPy, PyWavelets, and NumPy Python libraries were used to calculate features [61]–[63].

Additionally, participant demographics (Disease Status, Age, Gender, Weight, and Height) were merged from the study’s REDCap database. Exercise types (resistance, aerobic, breaks between exercises, or rest before exercise begins) were tagged during the experiments and synchronized with the matched timestamps. RPE values were joined using the nearest value within a one-minute tolerance; any data points without an RPE value within this window were excluded. Finally, any rows missing any of the described features were removed from the dataset. The full list of features is provided in Table 2. The total number of data points generated was 3,179.

**Table 2:** Summary of Extracted Features by Sensor Type.

| Sensor Type | Feature |  |
|---|---|---|
| Empatica Digital Biomarker | Pulse rate and actigraphy counts |  |
| Accelerometer [59] | **Mother Wavelet** |  |
|  | Daubechies 2 | [64] $\frac{1}{N}\sum_{j=6}^{7}\|cD_j^{Vert}\|^2$ and $\sum_{j=4}^{7}\|cD_j^{AP}\|^2/\|x^{AP}\|^2$ |
|  |  | $\|cD_1\|^2, \|cD_2\|^2, \|cD_3\|^2, \|cD_4\|^2,$ and $\|cD_5\|^2$ |
|  |  | $\|cD_1\|, \|cD_2\|, \|cD_3\|, \|cD_4\|,$ and $\|cD_5\|$ |
|  | Daubechies 3 | [65] $\|cD_4\|^2$ and $\|cD_5\|^2$ |
|  | Daubechies 4 | [66] Fractal Dimension = 2 − (*β* − 1)/2 |
|  | Daubechies 5 | [67] $\sum_{j=4}^{5}\|cD_j^{Vert}\|^2$ and $\sum_{j=4}^{5}\|cD_j^{AP}\|^2$ |
|  |  | [68] $\sum_{j=2}^{6}\|cD_j\|^2+\|dA_3\|^2+\|dD_3\|^2$ |
|  |  | Includes standard deviation and RMS of coefficients for all three components. |
|  | Mean and SD |  |
|  | Mean, SD, median, and 25<sup>*th*</sup> and 75<sup>*th*</sup> percentile |  |
|  | Mean low and mean rectified high pass filtered signals |  |
|  | Principal frequency |  |
|  | Spectral density |  |
|  | Mean DC, energy, entropy, and correlations between axes [69] |  |
|  | Magnitude of first five components of FFT analysis |  |
| Electrodermal Activity (EDA) [60] | Mean amplitude |  |
|  | Derived from first and second derivative | Maximum |
|  |  | Minimum |
|  |  | Maximum absolute value |
|  |  | Mean absolute value |
|  | Derived from wavelet coefficients | Maximum |
|  |  | Mean |
|  |  | Standard deviation |
|  |  | Median |
|  |  | Number above zero |
| Temperature | Mean |  |
|  | Standard deviation |  |
| Manually tagged | Exercise type |  |

**Note:** *N* represents the number of peaks in the coefficients of *cD*<sub>5</sub> as an approximation for steps taken; *x*<sup>*AP*</sup> represents the anterior-posterior acceleration; $cD_j^{Vert}$ and $cD_j^{AP}$ refer to the wavelet coefficients related to the vertical and anterior/posterior acceleration; *β* represents the slope of the variance of the detail coefficient plotted against decomposition level.
Table adapted in part from Table I and Table II in Preece et al. [59] and Table I in Taylor et al. [60].

#### Model Training and Cross-Validation

We implemented both classification and regression workflows using random forest models to predict RPE from the multi-modal sensor data using Scikit-learn [70]. Random forest models were selected based on our preliminary analysis comparing classification performance across models [71]. To evaluate model generalizability while limiting data leakage of the time series data, we used a 3-fold cross-validation scheme without shuffling. Each fold was stratified by RPE label and exercise type to ensure a consistent distribution of classes across training and testing sets.

Two primary modeling approaches were compared:

1. *PR-Only*: A univariate model using only pulse rate.
2. *Multi-Feature*: A multi-modal model incorporating all extracted physiological and inertial features.

#### Modeling Pipeline

A subject-specific pipeline was applied strictly within each training fold to prevent data leakage. To address the class imbalance inherent in subjective RPE reporting, training samples were weighted inversely proportional to their RPE frequency.

For the Multi-Feature models, initial feature ranking and dimensionality reduction were performed using Minimum Redundancy Maximum Relevance [72], [73]. This was followed by an iterative feature selection process where importance scores (mean impurity decrease) were derived from a baseline Random Forest model configured with 50 estimators. Rather than applying a fixed feature cutoff across all subjects, the optimal number of retained features was treated as a dynamically tunable hyperparameter.

Given the high inter-subject physiological heterogeneity in the ESKD cohort, particularly regarding fitness and medication regimens, model architectures were optimized individually rather than forcing a global hyperparameter consensus. A 3-fold cross-validation grid search was performed entirely within the training folds to determine the optimal pipeline configuration for each iteration. For Multi-Feature models, the grid search optimized the data scaling strategy, the number of top-ranked features selected, the number of Random Forest estimators, and the maximum tree depth. For PR-Only models, the grid search optimized the data scaling strategy, the number of estimators, and the maximum tree depth. Feature selection parameters were omitted because the model is univariate. This individualized tuning approach ensures that each model’s complexity and feature space are appropriately tailored to the participant’s unique autonomic and cardiovascular baseline.

#### Evaluation Metrics

Classification performance was quantified using balanced accuracy to mitigate the effects of non-uniform RPE distribution. Additionally, to better reflect the practical application of these models in a therapeutic setting, the RPE target variables were grouped into four distinct functional zones based on the Houston Methodist exercise prescription protocol for participants with ESKD: 0–2 (below prescribed exertion), 3– 5 (initial prescription target), 6–7 (progression and maintenance target, typically prescribed after initial adaptation), and 8–10 (above prescribed exertion, representing a potential safety risk). This provides a measure of clinical utility for real-time monitoring. For regression, models were evaluated using root mean squared error (RMSE); however, mean squared error (MSE) and mean absolute error (MAE) were also computed for comparison to other works.

#### Statistical Analysis

To evaluate the statistical significance of performance differences across our experimental conditions, two-way factorial Analyses of Variance (ANOVA) were conducted, treating each participant’s Multi-Feature and PR-Only results as separate observations, with the following factors:

- Model Type: Multi-Feature and PR-Only
- Disease Status: Control (no disability or chronic condition) and ESKD participants.

Prior to statistical testing, the assumption of normality for the residuals was verified using the Shapiro-Wilk test [74]. The raw RMSE residuals were not normally distributed, so a standard logarithmic transformation was applied to RMSE prior to ANOVA. The primary two-way ANOVAs were performed to examine the main and interaction effects of *model type* (Multi-Feature and PR-Only) and *disease status* (ESKD and Control) on both classification and regression performance. Following significant ANOVA findings, post-hoc pairwise comparisons were conducted using paired t-tests for within-subject comparisons and Welch’s independent t-tests for between-cohort comparisons, with *p*-values adjusted via the Holm-Bonferroni correction for multiple comparisons [75]. Because Holm-adjusted *p*-values are non-decreasing in the order of the raw *p*-values, some comparisons share the same adjusted value.

Subsequently, to investigate the influence of participant demographics (e.g., frailty status, age, BMI, gender) on regression performance (log-transformed RMSE), separate Linear Mixed Models (LMMs) were constructed for each individual demographic factor. LMMs were used for these variables because the data failed to meet the necessary assumptions for standard ANOVA. Data for activity type failed to meet necessary assumptions for standard ANOVA or LMM, therefore Generalized Estimating Equation (GEE) was used. For significant categorical effects identified by the individual LMMs and GEE, non-parametric Wilcoxon signed-rank tests (paired comparisons between model types) and Mann-Whitney U tests (unpaired comparisons) were used for post-hoc pairwise comparisons, with *p*-values adjusted within each factor using the Holm-Bonferroni correction [75]. Because these secondary analyses were exploratory, *p*-values for the LMM and GEE omnibus tests are reported without correction. All statistical analyses were evaluated at a significance level of *α* = 0.05.

#### Linear Regression

To further evaluate the relationship between heart rate and perceived exertion, a linear regression analysis was performed on heart rate (measured with a Polar H10 monitor) and reported RPE consistent with prior work by Borg and Kaijser [36]. Data were separated by disease status and ordinary least squares regression was performed. Heart rate measured by Polar H10 was used instead of pulse rate measured by the Empatica EmbracePlus to provide a more robust comparison with previous values reported by Borg and Kaijser [36].

## 3 Results

### 3.1 Pulse Rate Cannot Fully Predict RPE for ESKD

The multi-modal Multi-Feature set model consistently outperformed the PR-Only model for ESKD across both regression (*Model Type* main effect: *F*(1, 48) = 6.35, *p* = 0.015; paired t-test: *t* = −5.81, *p* = 0.0003) and classification (*Model Type* main effect: *F*(1, 48) = 4.85, *p* = 0.032; paired t-test: *t* = 3.19, *p* = 0.016) tasks (Fig. 1, Table 3). For the regression task, the ESKD Multi-Feature and PR-Only models achieved a mean Root Mean Squared Error (RMSE) of 1.33 (95% confidence interval (CI): 1.13–1.56) and 1.69 (CI: 1.44–1.98), respectively. For the classification task, the ESKD Multi-Feature and PR-Only models achieved a mean balanced accuracy of 48.6% (CI: 41.7%–55.6%) and 39.2% (CI: 32.3%–46.2%), respectively.

**Figure 1:**
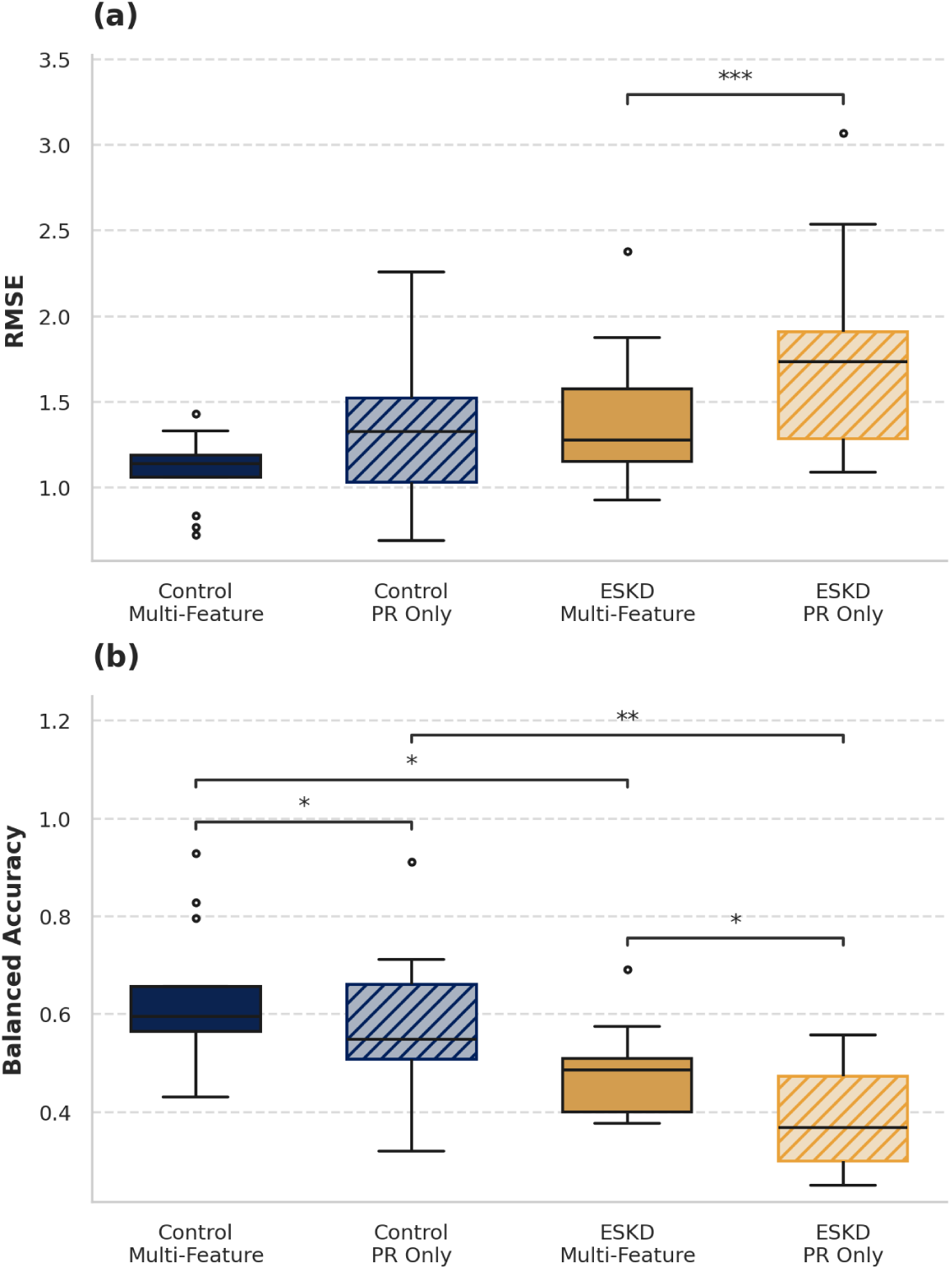
(a) RMSE of individually tuned regression models sorted by *Disease Status* and *Model Type*. (b) Unbinned balanced accuracy of individually tuned classification models sorted by *Disease Status* and *Model Type*. * *p* < 0.05, ** *p* < 0.01, *** *p* < 0.001

**Table 3:**
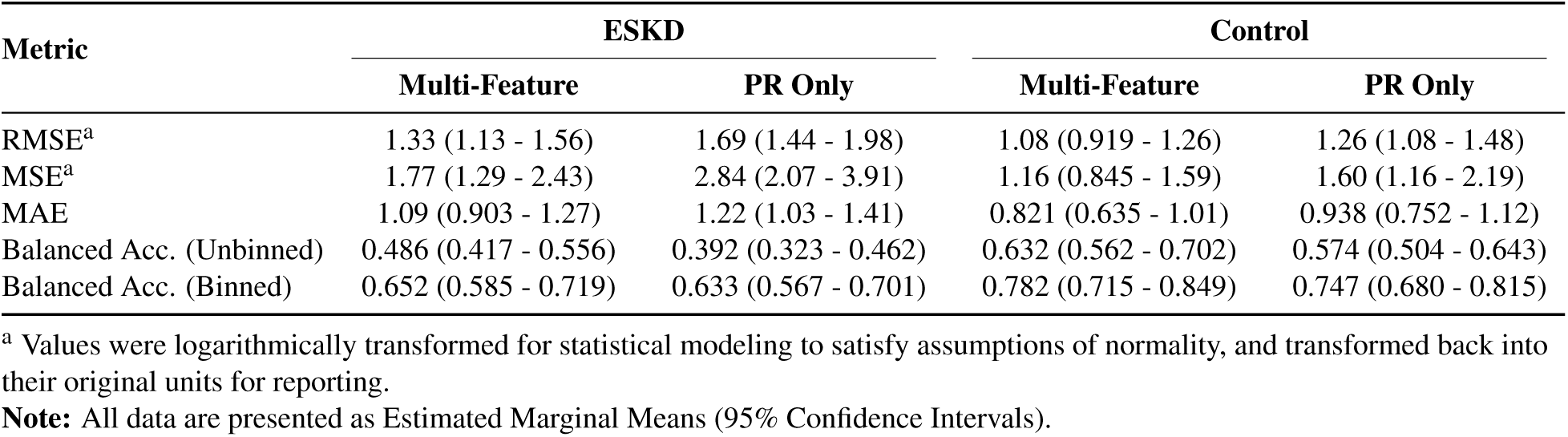
Summary of Model Performance.

To further elucidate pulse rate challenges in ESKD, we performed a linear regression analysis between RPE and heart rate and compared the fitted slope and correlation with those previously reported by Borg and Kaijser [36] (slope: *m* = 0.075; correlation: *r* = 0.881). The control participants exhibited a linear relationship between RPE and heart rate (*m* = 0.071; *r* = 0.741) which more closely approximated the values reported by Borg and Kaijser [36] than those of the ESKD participants (*m* = 0.026; *r* = 0.159) (Fig. 2). This suggests that heart rate is not modulated in ESKD participants or, consequently, their pulse rate despite changes in RPE.

**Figure 2:**
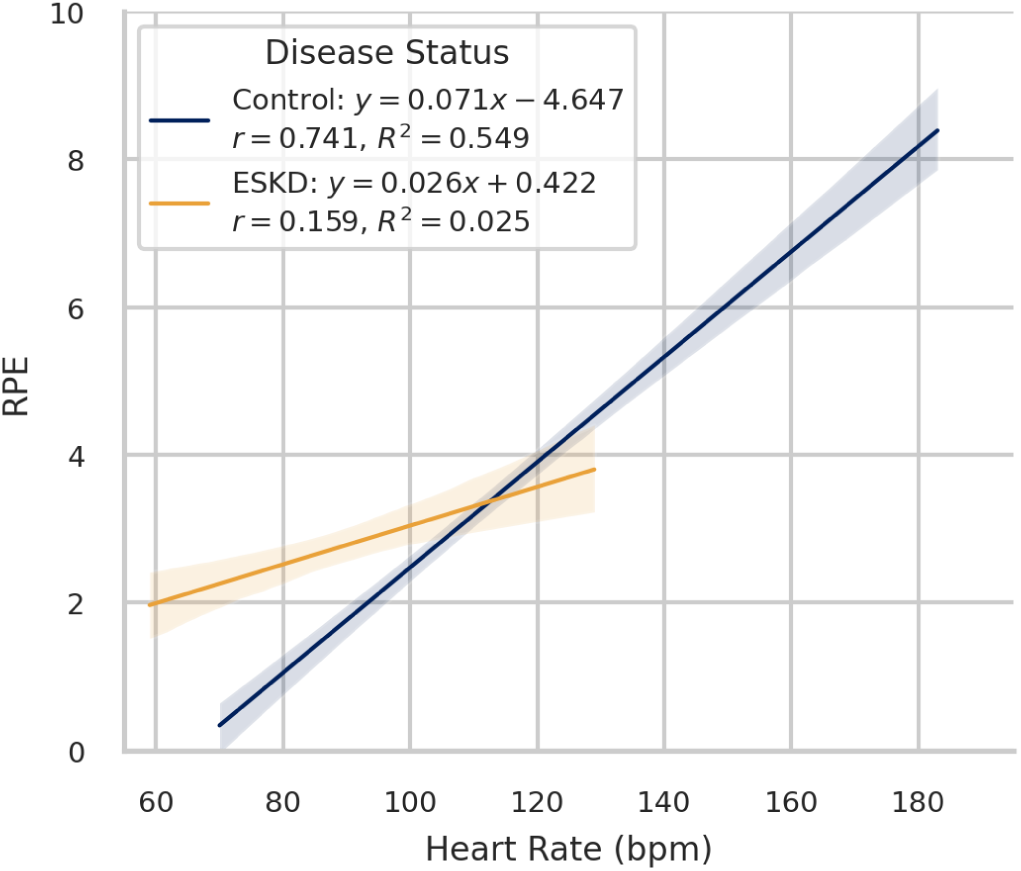
Linear regression of RPE vs heart rate measured by Polar H10 heart rate monitor. Data from ESKD participants are shown in orange and Control participants are shown in blue. Shading around regression lines represents the 95% confidence interval for the regression best fit line.

### 3.2 Classification Task Does Not Sufficiently Capture Variability in ESKD

We determined which task structure (regression versus classification) performed comparably to Control for ESKD by assessing the impact of *Disease Status* (ESKD versus Control) on performance metrics. Disease status had a significant main effect for the regression task (*F*(1, 48) = 10.1, *p* = 0.0026); however, post hoc comparisons did not show significant differences for ESKD and Controls for either *Model Type* (Multi-Feature independent t-test: *t* = −2.18, *p* = 0.073; PR-Only independent t-test: *t* = −2.32, *p* = 0.073). There were no significant interaction effects (*F*(1, 48) = 0.25, *p* = 0.62).

Disease status had a significant main effect for the classification task (*F*(1, 48) = 22.3, *p* < 0.001); Control outperformed ESKD for both Multi-Feature (*t* = 3.1, *p* = 0.012) and PR-Only (*t* = 3.5, *p* = 0.0076) models (Fig. 1, bottom, Table 3). Additionally, Multi-Feature outperformed PR-Only for Control participants (*t* = 2.69, *p* = 0.020). There were no significant interaction effects (*F*(1, 48) = 0.27, *p* = 0.61). We also classified participants into binned clinically relevant RPE zones (Table 3); model performance improved substantially compared to unbinned RPE predictions. The Multi-Feature models achieved a mean binned balanced accuracy of 78.2% (CI: 71.5–84.9%) for the Control group and 65.2% (CI: 58.5–71.9%) for the ESKD group. Disease status had a significant main effect (*F*(1, 48) = 13.3, *p* < 0.001; Multi-Feature independent t-test: *t* = −3.1, *p* = 0.011; PR-Only independent t-test: *t* = −2.2, *p* = 0.038). Model type had no significant effect (*F*(1, 48) = 0.62, *p* = 0.44), and there were no interaction effects (*F*(1, 48) = 0.058, *p* = 0.81).

Further analyses into confusion matrices generated from unbinned and binned classification tasks found that incorrect predictions tended to predict adjacent RPE predictions for both ESKD and Control (Fig. 3). However, incorrect predictions were closer to the correct labels for Control compared to ESKD on average (MAE Control: 0.67, CI: 0.478–0.852; MAE ESKD: 0.90, CI: 0.671–1.138). Our combined results suggest that classification performs worse for ESKD compared to Controls. Therefore, we focused on regression analyses for the remainder of the paper.

**Figure 3:**
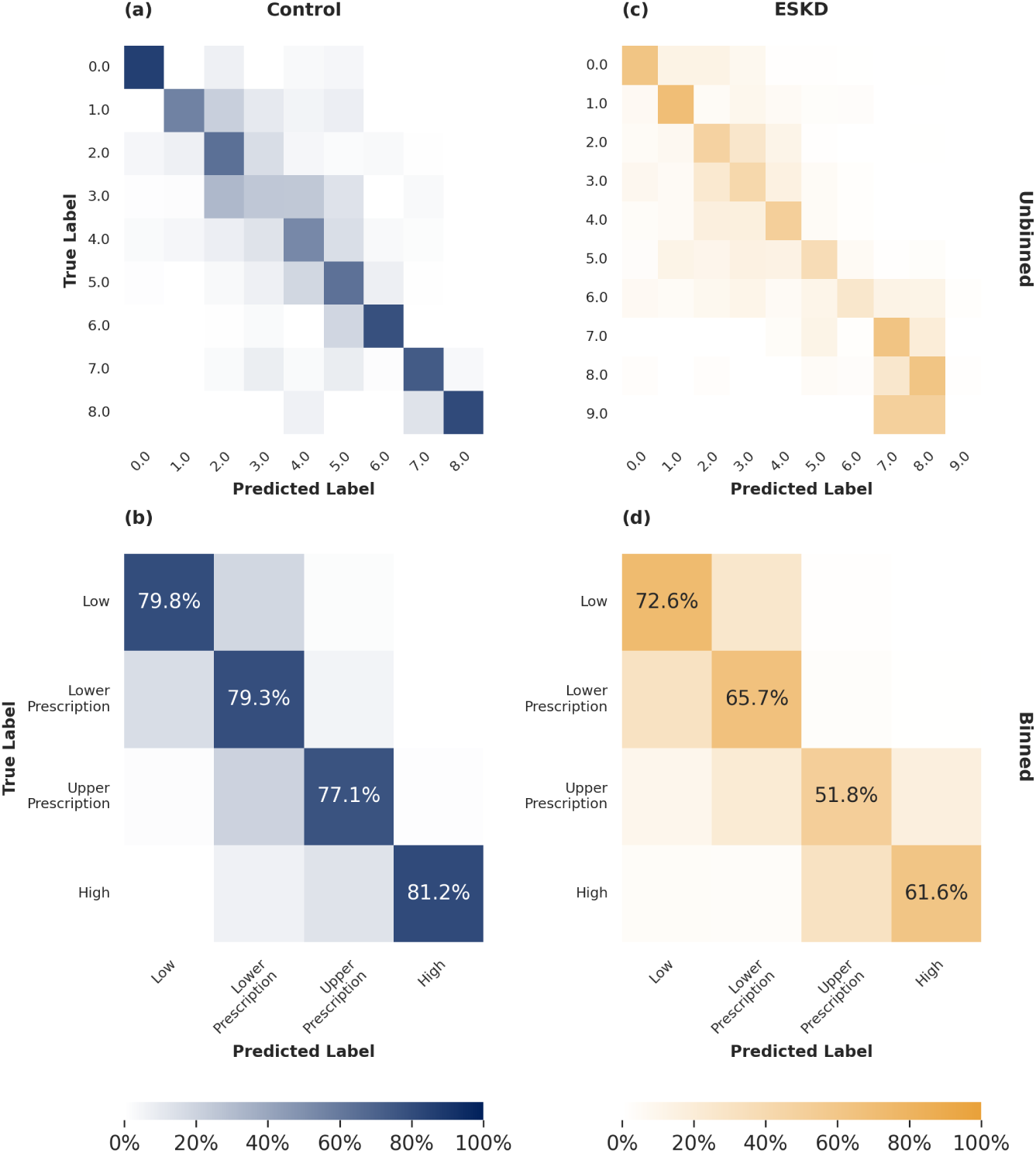
Averaged classification results for individually tuned random forest classifiers. For all figures, all data-points were predicted for each participant using Scikit-Learn’s cross_val_predict function [70], [76], and column percentages were calculated for each participant before being averaged across participants. (a) Classification predictions for control participants. (b) Classification predictions for control participants binned into clinically relevant categories as prescribed by Houston Methodist. (c) Classification predictions for ESKD participants. (d) Classification predictions for ESKD participants binned into clinically relevant categories as prescribed by Houston Methodist.

### 3.3 Important Regression Features for ESKD

The composition of the subject-specific regression models revealed distinct physiological dependencies based on disease status. For ESKD participants, the most prevalent features included mean skin temperature (92.3%), EDA mean (76.9%), the 25th percentile of x-axis acceleration (76.9%), EDA maximum (69.2%), and pulse rate (69.2%) (Table 4). However, high selection prevalence did not strictly correlate with high feature importance. Although mean temperature was the most frequently selected feature, it maintained a relatively low mean importance (Mean Decrease in Impurity [MDI]: 0.057 ± 0.0495). In contrast, maximum skin conductance, selected as the fourth most common, exerted a substantially greater impact on model output when present (MDI: 0.11 ± 0.210).

**Table 4:** Top 10 Features and Overall Feature Counts by Task and Group. Feature importance is measured as the Mean Decrease in Impurity (MDI).

|  | **ESKD** |  |  | **Control** |  |  |
|---|---|---|---|---|---|---|
| **Rank** | **Feature** | **Prev<br>(%)** | **Importance<br>(MDI ± SD)** | **Feature** | **Prev<br>(%)** | **Importance<br>(MDI ± SD)** |
| **Regression Models** |  |  |  |  |  |  |
| 1 | Mean Temperature | 92.3% | 0.057 ± 0.050 | **Pulse Rate** | 84.6% | 0.242 ± 0.289 |
| 2 | Mean EDA | 76.9% | 0.084 ± 0.163 | Mean EDA | 76.9% | 0.098 ± 0.193 |
| 3 | Acc X 25th Percentile | 76.9% | 0.018 ± 0.028 | Mean Temperature | 69.2% | 0.096 ± 0.226 |
| 4 | Max EDA | 69.2% | 0.112 ± 0.210 | Max EDA | 61.5% | 0.048 ± 0.076 |
| 5 | **Pulse Rate** | 69.2% | 0.094 ± 0.124 | EDA Wavelet L3 SD | 53.8% | 0.015 ± 0.030 |
| 6 | Acc Y 75th Percentile | 61.5% | 0.041 ± 0.095 | Acc Y Wavelet L1 SD | 46.2% | 0.023 ± 0.039 |
| 7 | Acc Z 75th Percentile | 61.5% | 0.025 ± 0.057 | Acc Z 25th Percentile | 46.2% | 0.022 ± 0.042 |
| 8 | Acc Z Fractal Dimension | 53.8% | 0.037 ± 0.116 | Acc X Wavelet L1 Mag. | 46.2% | 0.015 ± 0.036 |
| 9 | EDA Wavelet L1 Median | 53.8% | 0.031 ± 0.061 | Acc X Wavelet L2 SD | 46.2% | 0.012 ± 0.020 |
| 10 | Actigraphy Counts (X) | 53.8% | 0.026 ± 0.039 | Acc X Fractal Dimension | 46.2% | 0.010 ± 0.024 |
|  | *Total Features Used* |  | 29 ± 13.9 | *Total Features Used* |  | 21 ± 13.9 |
| **Classification Models** |  |  |  |  |  |  |
| 1 | Mean Temperature | 100.0% | 0.063 ± 0.031 | Acc Z Median | 92.3% | 0.033 ± 0.029 |
| 2 | **Pulse Rate** | 100.0% | 0.045 ± 0.027 | **Pulse Rate** | 84.6% | 0.063 ± 0.048 |
| 3 | Max EDA | 92.3% | 0.054 ± 0.030 | Max EDA | 76.9% | 0.046 ± 0.051 |
| 4 | Mean EDA | 92.3% | 0.051 ± 0.040 | Mean EDA | 76.9% | 0.038 ± 0.033 |
| 5 | Acc Z 25th Percentile | 84.6% | 0.025 ± 0.017 | Acc Z 75th Percentile | 76.9% | 0.021 ± 0.020 |
| 6 | Acc Y Median | 69.2% | 0.020 ± 0.019 | Acc Z 25th Percentile | 76.9% | 0.020 ± 0.017 |
| 7 | Acc Y Wavelet Energy | 69.2% | 0.017 ± 0.013 | Acc Z Mean | 69.2% | 0.024 ± 0.030 |
| 8 | Acc Z Wavelet Energy | 61.5% | 0.021 ± 0.024 | Actigraphy Counts (X) | 69.2% | 0.019 ± 0.024 |
| 9 | Acc X Mean | 61.5% | 0.021 ± 0.018 | Mean Temperature | 61.5% | 0.047 ± 0.052 |
| 10 | Acc Y 75th Percentile | 61.5% | 0.020 ± 0.019 | Acc X Wavelet L2 Mag. | 61.5% | 0.025 ± 0.033 |
|  | *Total Features Used* |  | 32 ± 4.5 | *Total Features Used* |  | 37 ± 7.0 |

*Note:* MDI = Mean Decrease in Impurity.

In the Control group, feature importance followed a more hierarchical structure, where importance generally scaled with selection prevalence. Pulse rate was the dominant predictor for control participants, appearing in 84.6% of models with a mean importance of 0.24, more than double its importance in the ESKD group (0.094). Other prevalent features for Controls included mean skin conductance (76.9%), mean temperature (69.2%), maximum skin conductance (61.5%), and the standard deviation of the L3 wavelet for EDA (53.8%). Notably, ESKD regression models required more features, using an average of 29 ± 13.9 features per subject compared to 21 ± 13.9 features for the Control group (Table 4).

### 3.4 Analysis of Confounding Features

To assess the impact of *Model Type* (Multi-Feature, PR-Only) and potentially confounding demographic variables, frailty status, age, BMI, and gender were separately evaluated across the combined ESKD and Control cohorts using linear mixed models (LMM). There were no main or interaction effects for any of the demographic variables of interest (*p* > 0.05). There were *Model Type* main effects for frailty status (*χ*^2^(1, *N_total_* = 52) = 4.4, *p* = 0.037), age (*χ*^2^(1, *N_total_* = 52) = 15.3, *p* = 0.0001), and gender (*χ*^2^(1, *N_total_* = 52) = 11.7, *p* = 0.0006). Post hoc Wilcoxon Signed-Rank tests demonstrated that Multi-Feature outperformed PR-Only for individuals who were prefrail (*W* = 0, *p* = 0.0059), < 65 years of age (*W* = 0, *p* = 0.0078), ≥ 65 years of age (*W* = 11, *p* = 0.0008), female (*W* = 7, *p* = 0.0029), and male (*W* = 2, *p* = 0.0029) (Fig. 4a–d).

**Figure 4:**
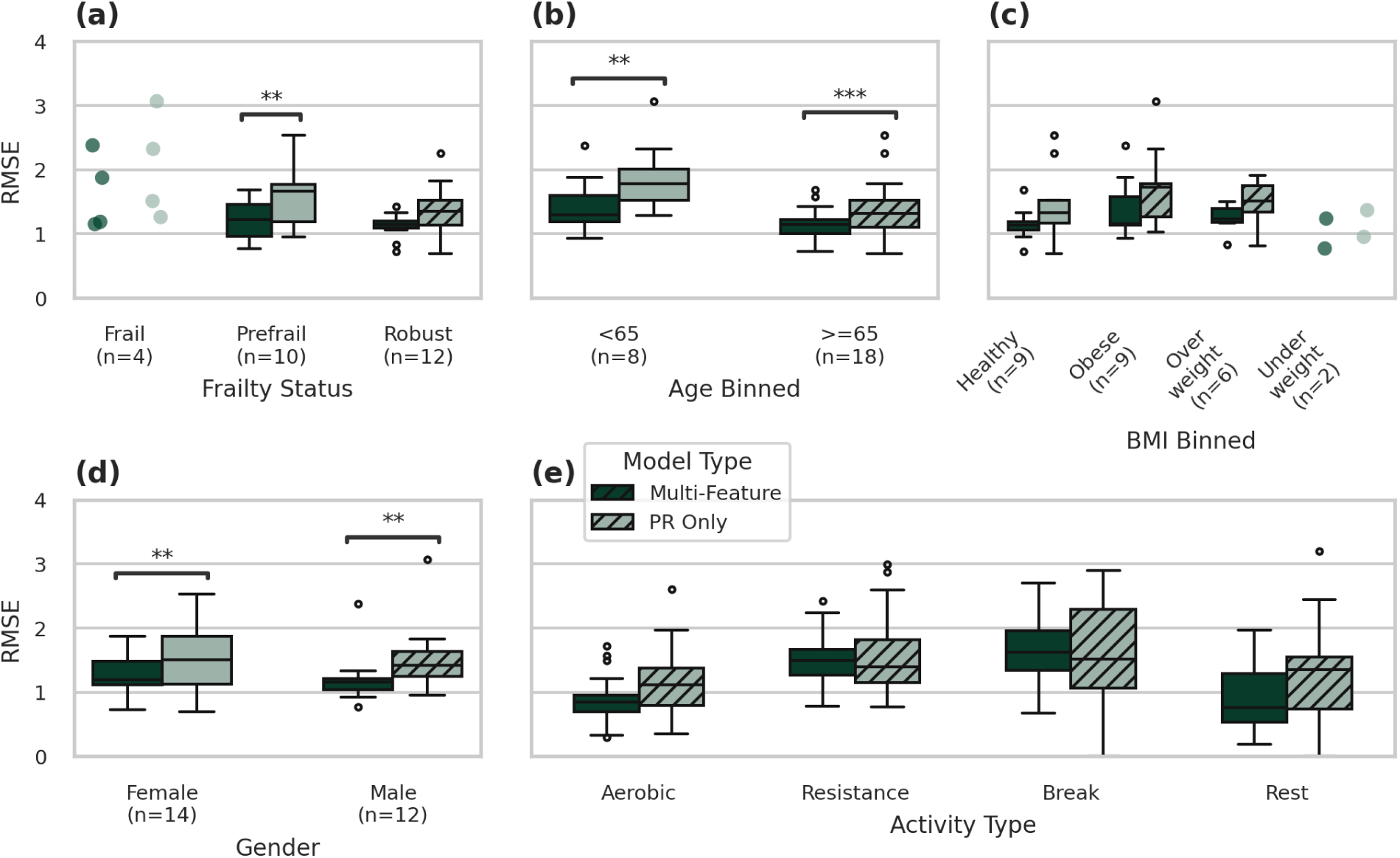
Performance of individually tuned regression models based on demographic features or activity type using root mean squared error (RMSE) and model type (Multi-Feature models and pulse rate (PR) only models). ESKD and control participant data were pooled together due to the low number of participants. (a) Performance by frailty status. Model type was significant for prefrail participants. (b) Performance by age. Model type was significant for participants of both age groups (below 65 years of age and 65 years of age or older). (c) Performance by body mass index (BMI). No significant main effects or interactions. (d) Performance by gender. Model type was significant for both male and female participants. (e) Performance by activity type. Activity types were aerobic exercise, resistance exercise, breaks between sets, and rest at the beginning of the session. No statistical significance determined by post hoc tests. Significance for (a)-(d) was first determined using LMM, and (e) used GEE. All reported significance was calculated using Mann-Whitney U Tests or Wilcoxon Signed-Rank Tests. * *p* < 0.05, ** *p* < 0.01, *** *p* ≤ 0.001

Furthermore, a Generalized Estimating Equation (GEE) analysis revealed significant main effects (*Model Type*: *χ*^2^(1), *p* < 0.0001; activity type: *χ*^2^(3), *p* < 0.0001) and an interaction effect between *Model Type* and *Activity Type* (aerobic, resistance, breaks between sets, and rest before exercise begins) (*χ*^2^(3), *p* = 0.0001). However, subsequent post hoc Mann-Whitney U and Wilcoxon Signed-Rank tests did not identify any statistically significant differences between specific subgroups (Fig. 4e).

## 4 Discussion

### 4.1 Beyond Pulse Rate for Predicting RPE in ESKD

In this work, we identified distinct differences between RPE prediction models tuned for ESKD participants and those tuned for Controls. For regression tasks, baseline models that relied solely on pulse rate to predict RPE performed substantially worse for ESKD participants than their Multi-Feature counterparts (*p* = 0.0003; RMSE: 1.33 and 1.69, respectively). In comparison, no significant performance degradation was observed when Controls were restricted to pulse rate alone. This contrast suggests that multi-modal features may be necessary for predicting exertion in ESKD.

The lower performance of PR-Only models can likely be explained in part by our linear regression analysis, where Controls had a larger slope for RPE versus heart rate than the ESKD group (*m* = 0.071 and *m* = 0.026, respectively), as well as a stronger correlation between the fitted line and experimental values (*r* = 0.741 and *r* = 0.159, respectively). The relationship for Controls was similar to that found in Borg and Kaijser [36] (*m* = 0.075, *r* = 0.881). The clear linear relationship between heart rate and RPE for the Controls should make their data more separable compared to the ESKD group, which lacked a clear linear relationship.

The lack of a relationship between heart rate and RPE for ESKD participants is expected, as frail individuals frequently exhibit impaired autonomic nervous system function, leading to smaller heart rate changes in response to physiological exertion [30]. Likewise, ESKD patients are commonly prescribed beta-blockers to control their blood pressure [32], which lowers their baseline heart rate and limits its ability to rise during exercise [31], [33]. In our study, 11 out of 13 ESKD participants were prescribed beta-blockers, and one of the two not taking beta-blockers was prescribed diltiazem, another heart-rate-reducing medication used to manage hypertension [77]. This may explain why the four most common features for ESKD participants were related to EDA, skin temperature, and movement, whereas pulse rate was only the fifth most common feature. This further suggests that multi-modal sensing is necessary to accurately capture exertion in ESKD patients.

Lastly, secondary analyses further support that the need for multi-modal sensing in the ESKD group was not driven by the participants’ demographics. The Multi-Feature models significantly outperformed the PR-Only models across both age groups (< 65: *p* = 0.0078, ≥ 65: *p* = 0.0008) and sexes (Female: *p* = 0.0029, Male: *p* = 0.0029), whereas no significant differences in model performance were observed by BMI or activity type. Because the Multi-Feature model’s advantage was consistent across age and sex rather than confined to a single demographic subset, it is unlikely that the performance differences for ESKD participants in our primary *Disease Status* and *Model Type* analysis are due to these characteristics. Furthermore, for robust participants (i.e., not frail or at risk of becoming frail according to the FFP), PR-Only models performed comparably to Multi-Feature models (*p* = 0.054). In contrast, Multi-Feature models significantly outperformed PR-Only models for prefrail participants (*p* = 0.0059). This reinforces the idea that underlying disease and early frailty status drive the necessity for multi-modal exertion tracking.

### 4.2 Regression Performs Comparably to Prior Work

Comparing predictive performance to existing literature is challenging due to the heterogeneity of methodologies, study populations, and exercise protocols. Nevertheless, the performance of our Multi-Feature Control models aligns closely with, and in several cases exceeds, previously reported benchmarks for regression tasks. Our regression outcome measures for Controls (MAE [42], [43], [45], RMSE [45], [53], MSE [41]) performed comparably to prior work predicting RPE from wearable sensor data. For example, Kim et al. [45] and Chowdhury et al. [53] reported RMSE values of 1.21 and 1.84, respectively, comparable to the RMSE of 1.08 (95% CI: 0.919–1.26) that we observed. This is notable because these prior works do not use sensors suitable for ESKD patients undergoing long-term prehabilitation (e.g., IMU systems, chest-strap heart rate monitors, etc.), and all works except Ibrahim et al. [49] include young, healthy adults. One notable exception was Gholami et al. [38], who achieved an RMSE value of 0.22, significantly lower than the RMSE of 1.08 (95% CI: 0.919–1.26) we obtained. However, their work predicts RPE in young, healthy female athletes performing only a running task using multiple custom lower-body joint angle sensors, whereas our work is with older adults and incorporates both resistance and aerobic exercises. Predicting across exercise types introduces more variance into the accelerometer-based features, so lower performance is not unexpected.

Upon initial consideration, our classification models achieved a binned balanced accuracy of 78.2%, which is lower than or comparable to the raw accuracies of 74–98% reported in prior studies [41], [47], [48], [78]. However, a direct comparison requires careful consideration of the specific validation frameworks and target definitions employed. For example, some studies used binary classification schemes with a chance accuracy of 50% [41], [47], [48], whereas this work uses multiclass binned RPE with a chance accuracy of 25%. The lower number of classes and higher chance accuracy will inherently increase accuracy scores. Additionally, although Pinto-Bernal et al. [78] reported a 98% accuracy using a binned RPE approach, their methodology involved randomly shuffling data to divide it into training and testing folds. In prior work, we also achieved higher classification accuracies when using random assignment to test and train folds [71].

### 4.3 Critical Analysis of Results

Although this study establishes the necessary preliminary evidence that ESKD patients exhibit fundamentally altered physiological signals that require different features than those of Controls, several limitations must be acknowledged. Primarily, the study was constrained by a relatively small sample size and was limited to a single exercise session per participant. Although we employed rigorous cross-validation with unshuffled, stratified splits to mitigate data leakage and promote model robustness, relying on a single session introduces a minor, unavoidable degree of leakage across folds. Addressing this by strictly bisecting the session data was not methodologically viable given the structure of the prehabilitation protocol; a halfway split would completely segregate aerobic and resistance exercises into separate datasets, leading to highly imbalanced feature spaces and unrepresentative RPE distributions. Furthermore, requiring this vulnerable patient population to attend multiple preliminary data collection sessions would have been unduly burden-some at this early pilot stage of research. Our models serve as an exploratory baseline identifying which sensor modalities are strictly necessary for this population. Future validation on longitudinal, inter-session data is an important next step.

Additionally, the restricted distribution of the target RPE variable limits the generalizability of our models at maximal exertion. While low-to-medium exertion levels are well represented in our dataset, RPE values at the highest end of the scale (i.e., 8–10) are sparse. Inducing near-maximal or maximal exertion poses an unjustifiable safety risk to older adult Controls and a severe cardiovascular risk to the ESKD cohort. Consequently, our models are optimized for tracking mild-to-moderate prehabilitation activities rather than peak physical exertion. Despite these limitations, our findings provide a foundation for remote exertion monitoring in ESKD prehabilitation, demonstrating that clinical deployment of wearable technologies will require condition-specific modeling architectures.

### 4.4 Future Directions

Future research should expand upon these findings by incorporating larger cohorts and multiple in-person prehabilitation sessions. Expanding the protocol to at least two sessions per participant will substantially enhance the reliability of the results by enabling the evaluation of subject-specific models on completely unseen, temporally distinct holdout data. Furthermore, longitudinal data collection will naturally increase the probability of capturing physiological signals at higher exertion levels; although participants were not explicitly instructed to push into high-intensity zones, it was not uncommon for individuals to transiently report RPE values above 7 during the protocol. Inclusion of more of these exertion spikes in future datasets will contribute to training robust models capable of safely recognizing maximal exertion boundaries.

Once these preliminary models are validated on larger, multi-session datasets, the predictive pipelines should be optimized for real-time inference and integrated into mobile health platforms. This translation will allow researchers to test the reliability of wearable-based RPE prediction in unstructured, unsupervised home settings. Ultimately, deploying these validated algorithms will provide an objective measure of both exertion and adherence, enabling rigorous evaluation of home-based exercise interventions aimed at reducing frailty. Beyond ESKD, this analytical framework should be expanded to investigate other chronic diseases and clinical populations whose unique physiological profiles are systematically underserved by commercial fitness algorithms trained predominantly on healthy adults.

## 5 Conclusion

This study demonstrates that multi-modal commercial smartwatches can effectively predict the Rating of Perceived Exertion (RPE) in frail and prefrail patients with End-Stage Kidney Disease (ESKD) during prehabilitation. Our findings confirm that models relying solely on pulse rate—the current clinical standard for exercise prescription—are insufficient for this population given their altered physiological responses and prevalent beta-blocker use. By employing individualized machine learning pipelines, we showed that multi-modal models using electrodermal activity, skin temperature, and accelerometer data significantly outperform pulse-rate-only models for ESKD patients. Furthermore, the differences in feature importance and selection prevalence between the ESKD and healthy Control cohorts highlight that predictive algorithms trained on healthy adults cannot be directly mapped to chronically ill populations. While limited by a single-session protocol and a small sample size, this work establishes a foundation for remote, wearable-based exertion monitoring. Future work must validate these individualized models across longitudinal, multi-session datasets to translate this framework into a robust, real-time tool for safely monitoring and progressing home-based ESKD prehabilitation.

## Declarations

### Ethics approval and consent to participate

This study was approved by the Institutional Review Boards of Rice University (IRB-FY2024-244) and Houston Methodist Hospital (PRO00038084:1). Written informed consent was obtained from all participants prior to the start of the study sessions.

### Clinical trial registration

The portion of the study including ESKD patients is registered at ClinicalTrials.gov (NCT06886256).

### Data availability

The data underlying this study contain participant health information and are not publicly available. De-identified data may be made available from the corresponding author upon reasonable request, subject to approval by the Rice University and Houston Methodist Hospital Institutional Review Boards.

## Acknowledgments

The authors would like to thank all of the study participants for their invaluable time, effort, and commitment to this research. We also express our gratitude to Zheng Xiao for his contributions to the technical infrastructure and data collection support for this study.

